# Defining tumor growth in vestibular schwannomas: a volumetric inter-observer variability study in contrast-enhanced T1-weighted MRI

**DOI:** 10.1101/2024.03.15.24304080

**Authors:** Stefan Cornelissen, Sammy M. Schouten, Patrick P.J.H. Langenhuizen, Suan Te Lie, Henricus P.M. Kunst, Peter H.N. de With, Jeroen B. Verheul

## Abstract

For patients with vestibular schwannomas (VS), a conservative observational approach is increasingly used. Therefore, the need for accurate and reliable volumetric tumor monitoring is important. Currently, a volumetric cutoff of 20% increase in tumor volume is widely used to define tumor growth in VS. The goal of this study is to investigate the tumor volume dependency on the limits of agreement (LoA) for volumetric measurements of VS by means of an inter-observer study.

This retrospective study included 100 VS patients who underwent contrast-enhanced T1-weighted MRI. Five observers volumetrically annotated the images. Observer agreement and reliability was measured using the LoA, estimated using the limits of agreement with the mean (LOAM) method, and the intraclass correlation coefficient (ICC). Influence of imaging parameters and tumor characteristics were assessed using univariable and multivariable linear regression analysis.

The 100 patients had an average median tumor volume of 903 mm^3^ (IQR: 193-3101). Peritumoral cysts were found in 6 (6%) patients. Patients were divided into four volumetric size categories based on tumor volume quartile. The smallest tumor volume quartile showed a LOAM relative to the mean of 26.8% (95% CI: 23.7, 33.6), whereas for the largest tumor volume quartile this figure was found to be 7.3% (95% CI: 6.5, 9.7) and when excluding peritumoral cysts: 4.8% (95% CI: 4.2, 6.2). Of all imaging parameters and tumor characteristics, only tumor volume was associated with the LoA (adjusted B=-0.001 [95% CI: -0.001, 0.000; *P*=0.003]).

Agreement limits within volumetric annotation of VS are affected by tumor volume, since the LoA improves with increasing tumor volume. As a result, for tumors larger than 200 mm^3^, growth can reliably be detected at an earlier stage, compared to the currently widely used cutoff of 20%. However, for very small tumors, growth should be assessed with higher agreement limits than previously thought.

## 1 Introduction

Vestibular schwannomas (VS) are uncommon benign intracranial tumors that emerge from Schwann cells of the vestibulocochlear nerve. Management modalities consist of microsurgery, radiosurgery or observation. In the last decades, the focus of VS management has shifted from total surgical resection, with the inherent risk to neighboring neurovascular structures, to functional preservation^1–5^. The share of patients undergoing microsurgery is therefore decreasing and tumor observation (also called wait-and-scan) is becoming a common management strategy^3^. Tumor observation is particularly employed for smaller tumors, as growth is not always observed in these tumors and symptoms usually do not improve after treatment^3,6,7^. For larger tumors, microsurgery remains the most common treatment and management strategy. For smaller tumors that exhibit growth, the less invasive stereotactic radiosurgery is increasingly used with the goal of stabilizing these tumors^2,4,8^.

Because of this trend towards observation and the use of radiosurgery in VS management, the importance of accurate and reliable tumor monitoring is increasing. Within a wait-and-scan approach, correctly differentiating stable and growing tumors is relevant for clinical decision-making. The same applies for accurately observing tumor response after radiosurgery to define successful treatment, that is, the halt of further tumor growth^2^. Studies have shown that volumetric annotations are less prone to error compared to linear tumor monitoring and therefore provide the most robust results in VS tumor monitoring^9–13^. However, the accuracy and reliability in volumetric measurements are also subject to inter-observer variability. Several studies have been conducted to quantify this error source by defining the limits of agreement (LoA) between observers^9–11,14,15^. Based on these results, the current general consensus for the volumetric cutoff on VS tumor growth has been set on 20% between two sequential observations^2,11,12,15^.

However, applying a cutoff of 20% for volumetric analyses of all VS may be incorrect. It has already been implied that the LoA depends on tumor volume^11^. For that reason, the current general cutoff definition should be reappraised, in order to improve the reliability and robustness of volumetric VS tumor assessment and growth classification. This study investigates the tumor volume dependency of the LoA for volumetric measurements by means of an inter-observer study.

## 2 Methods

This retrospective study was conducted at Elisabeth-TweeSteden Hospital in Tilburg, the Netherlands, a tertiary referral hospital for VS microsurgery and Gamma Knife radiosurgery. Institutional review board approval was obtained and the requirement for informed consent was waived.

### 2.1 Materials

Elisabeth-TweeSteden Hospital has an established extensive database (including follow-up) of patients with unilateral sporadic VS treated with Gamma Knife radiosurgery^16,17^. All tumors were annotated and volumetrically analyzed using the Gamma Knife treatment planning software (GammaPlan, Version 11, Elekta AB, Stockholm, Sweden).

A total of 100 patients were selected randomly from the database, based on the natural distribution of VS tumor volumes in both wait-and-scan^7^ and radiosurgery cohorts^17^, see Figure 1. In order to verify whether this annotation dataset provides adequate statistical power, a required sample size was determined by the confidence interval (CI) lower limit procedure, which has proven to be a valid power analysis method in inter-observer reliability studies^18^. This method yielded a minimum required sample size of 26 (ρ=0.9, ρ_0_=0.8, β=0.8, α=0.05, N=5). This shows that the inclusion of 100 patients for the annotation dataset is sufficient.

**Figure 1:**
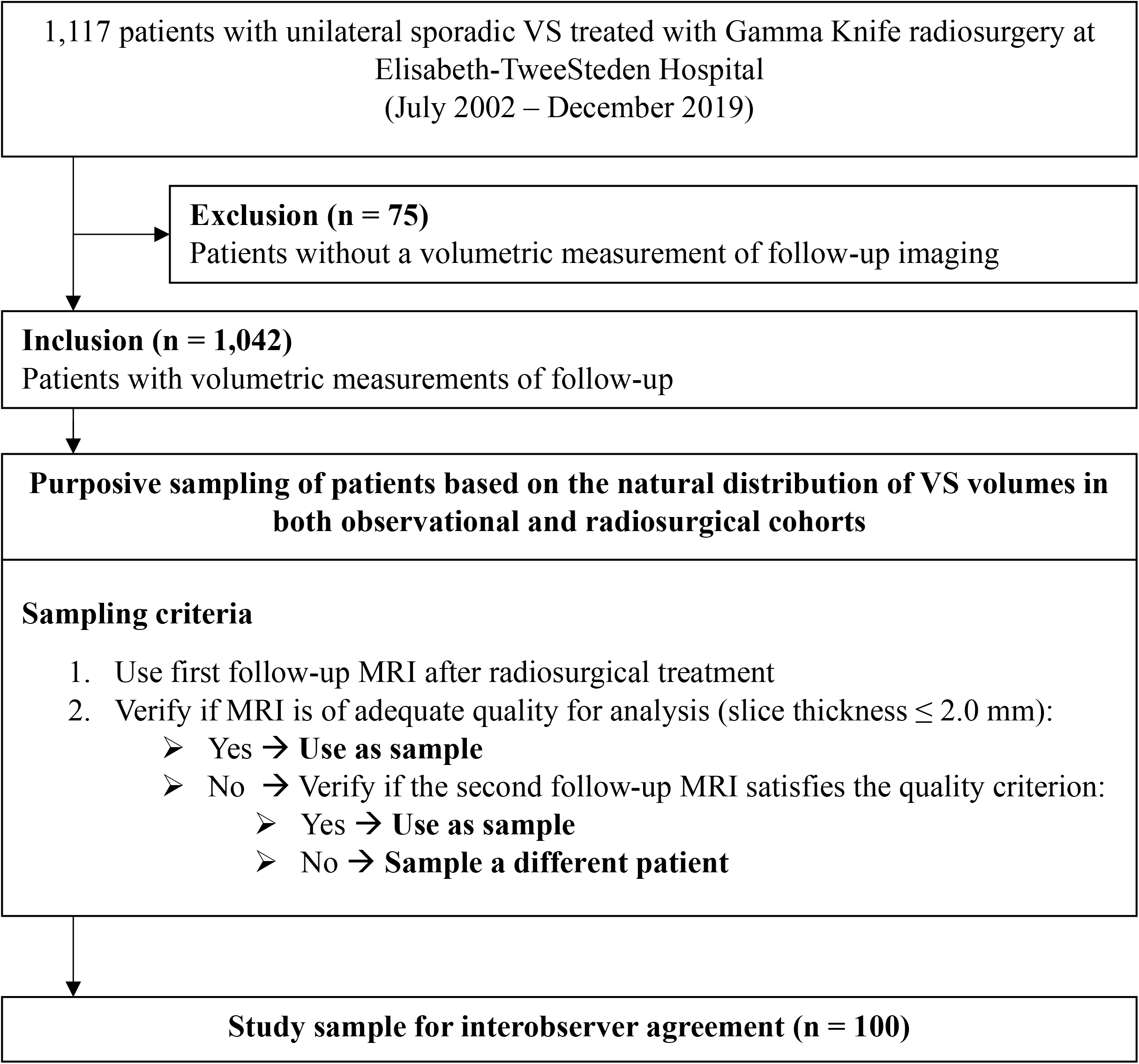
Flowchart showing the inclusion process

### 2.2 Imaging parameters

The patients included in our study underwent contrast-enhanced T1-weighted MRI, as part of their follow-up protocol after radiosurgical treatment. These scans were obtained between 2005 and 2020. Imaging was performed on the axial plane with either a 1.0 T, 1.5 T, or 3.0 T scanner (Achieva, Intera, and Ingenia; all Philips Healthcare, Best, The Netherlands). Gadoterate meglumine (Dotarem, Guerbet) was administered intravenously (5 to 10 mmol, depending on body weight). Image acquisition parameters varied throughout the years: median echo time 4.6 msec (range: 3.9 – 6.9), median repetition time 25 msec (range: 8.4 – 26.6), median slice thickness 1.6mm (range: 0.8 – 2.0), and median voxel spacing 0.78 mm (range: 0.25 – 1.0).

### 2.3 Observers and annotation

Five observers participated in this study: two senior neurosurgeons, both with extensive experience in segmenting VS as part of radiosurgical treatment planning, and three researchers with experience in segmenting VS as part of follow-up analyses. Annotation was performed in GammaPlan, occurred independently, while no prior information was available to the observer (e.g. earlier annotations or measurements). The semi-automated segmentation method included in GammaPlan aided the observers in segmenting the tumors. This method enables the observer to select a voxel-value range that corresponds to the voxel values of the tumor, resulting in a coarse initial segmentation. Following this, the observer can manually fine-tune the segmentation.

### 2.4 Statistical analyses

The average observed volume was calculated for each subject. Relative volume standard deviation (*SD*_*V*%_) was calculated by dividing the standard deviation of the observed volumes for each subject by its average observed volume. This metric is high when the observed volumes within a single subject substantially deviate between observers.

Observer agreement was assessed using the limits of agreement with the mean (LOAM) method^19^. This procedure is a generalization of the commonly used Bland-Altman plots, which calculates agreement limits for two observers. The LOAM method can be used for multiple observers and expresses the agreement limits as a confidence interval. More specific, this method calculates how much an individual measurement may ostensibly deviate from the mean of the measurements of all observers for a single subject. The method also allows for the detection of the source of the variation between observers by estimating both the inter-observer variance 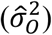, which represents the systematic differences between observers, and the residual variance 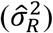, which represents the random measurement error. The intraclass correlation coefficient (ICC) follows from this computation as well.

Tumors were categorized into four size categories (I to IV) based on their respective volume quartiles. The LOAM and its variance components were calculated for each of the four size categories. Furthermore, in order to extend this analysis to the entire dataset, a sliding window of width 26 (i.e. our required sample size) was employed. Starting with the smallest 26 tumors, the LOAM and overall average observed volume were calculated for each set of 26 consecutive tumors, ending with the largest 26 tumors. Through this, it enabled us to calculate a volume-dependent continuous LOAM.

As we hypothesized based on clinical practice, that peritumoral cysts have an important effect on the inter-observer variability, we performed all analyses both including and excluding tumors with peritumoral cysts.

Both univariable and multivariable linear regression analyses were executed for all imaging parameters and tumor characteristics, in order to investigate any significant associations on the relative volume standard deviation (*SD*_*V*%_).

We considered *P* values smaller than 0.05 to indicate statistical significance. Analyses were performed by using statistical software (SPSS Version 27.0; SPSS, Chicago, Illinois) and in Python (Version 3.8.8).

## 3 Results

### 3.1 Patient characteristics

A total of 100 VS patients were included with a median tumor volume of 895 mm^3^ (IQR 214 – 3066), based on the earlier conducted volumetric analyses in our database. The study population had a median age of 58 (IQR: 50 – 67) at the time of follow-up. The distribution of tumor laterality was found to be almost equal for left (47) and right (53). Peritumoral cystic components were identified in 6 patients, which were all belonging to the two highest volume quartiles. Table 1 further outlines the patient and tumor characteristics in the annotation dataset.

**Table 1.** Patient and tumor characteristics in volume quartiles I–IV.

| Tumor type | N | Median average volume (mm<sup>3</sup>)* | Laterality (L/R)<sup>†</sup> | Peritumoral cystic component<sup>†</sup> | Age at Follow-Up* | Sex<sup>†</sup> |  |
| --- | --- | --- | --- | --- | --- | --- | --- |
|  |  |  |  |  |  | M | F |
| **All tumors** | 100 | 903 (193-3,101) | 47/53 (47/53) | 6 (6) | 58 (50-67) | 59 (59) | 41 (41) |
| I | 25 | 109 (63-138) | 15/10 (60/40) | 0 (0) | 64 (51-70) | 11 (44) | 14 (56) |
| II | 25 | 419 (291-594) | 8/17 (32/68) | 0 (0) | 59 (50-65) | 18 (72) | 7 (28) |
| III | 25 | 1,411 (1,132-2,198) | 9/16 (36/64) | 2 (8) | 57 (49-65) | 15 (60) | 10 (40) |
| IV | 25 | 5,359 (4,370-8,274) | 15/10 (60/40) | 4 (16) | 55 (50-69) | 15 (60) | 10 (40) |
| **Tumors without peritumoral cystic components** | 94 | 743 (175-2,376) | 43/51 (46/54) | 0 (0) | 58 (50-67) | 55 (59) | 39 (41) |
| **Tumors with peritumoral cystic components** | 6 | 7,030 (4,555-11,125) | 4/2 (67/33) | 6 (100) | 60 (50-70) | 4 (67) | 2 (33) |

Note.
\* Data in parentheses are inter-quartile range
<sup>†</sup> Data in parentheses are percentages

### 3.2 Agreement limits stratified by volume

The resulting figures after calculating the mean volume, LOAM, variance components, and the ICC are summarized in Table 2 for each individual size category. The results of the same analysis while excluding tumors with peripheral cysts, are presented in the same table. The corresponding agreement plots for all tumors are presented in Figure 2.

**Table 2.** LOAM, estimated variance decomposition (inter-observer 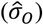, and residual 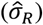), and ICC for all observers for all tumors (n = 100) and excluding tumors with peritumoral cysts (n = 94) divided into quartiles based on tumor volume

| | Mean<br>(mm<sup>3</sup>) | LOAM (mm<sup>3</sup>);<br>percent of mean volume (%) | $\hat{\sigma}_O$<br>(mm<sup>3</sup>) | $\hat{\sigma}_R$<br>(mm<sup>3</sup>) | ICC<br>(-) |
| --- | --- | --- | --- | --- | --- |
| **All tumors** |  |  |  |  |  |
| **I** | 100 | ± 27 (24-34);<br>26.8% (23.7-33.6) | 2 (0-7) | 15 (13-18) | .911 (.849-.954) |
| **II** | 468 | ± 49 (43-77);<br>10.4% (9.2-16.4) | 11 (2-21) | 25 (22-30) | .981 (.965-.991) |
| **III** | 1,655 | ± 126 (112-157);<br>7.6% (6.7-9.4) | 10 (0-31) | 63 (56-71) | .987 (.977-.994) |
| **IV** | 6,941 | ± 509 (450-677);<br>7.3% (6.5-9.7) | 70 (0-150) | 282 (247-328) | .995 (.991-.998) |
| **Excluding tumors with peritumoral cysts\*** |  |  |  |  |  |
| **III** | 1,606 | ± 106 (93-128);<br>6.6% (5.8-7.9) | 0<sup>†</sup> | 60 (53-70) | .990 (.981-.995) |
| **IV** | 6,305 | ± 302 (263-390);<br>4.8% (4.2-6.2) | 28 (0-84) | 170 (147-202) | .998 (.995-.999) |

**Note.** – Data in parentheses are 95% confidence intervals.
LOAM = limits of agreement with the mean, ICC = intra-class correlation coefficient.
\* Quartiles I and II do not contain any tumors with peritumoral cysts (see Table 1)
<sup>†</sup> Estimated variance was found to be negative. This may indicate that the true variance is near-zero, since the observed ICC shows excellent reliability. Consequently, this variance can only be accurately estimated by the inclusion of more observers.

**Figure 2:**
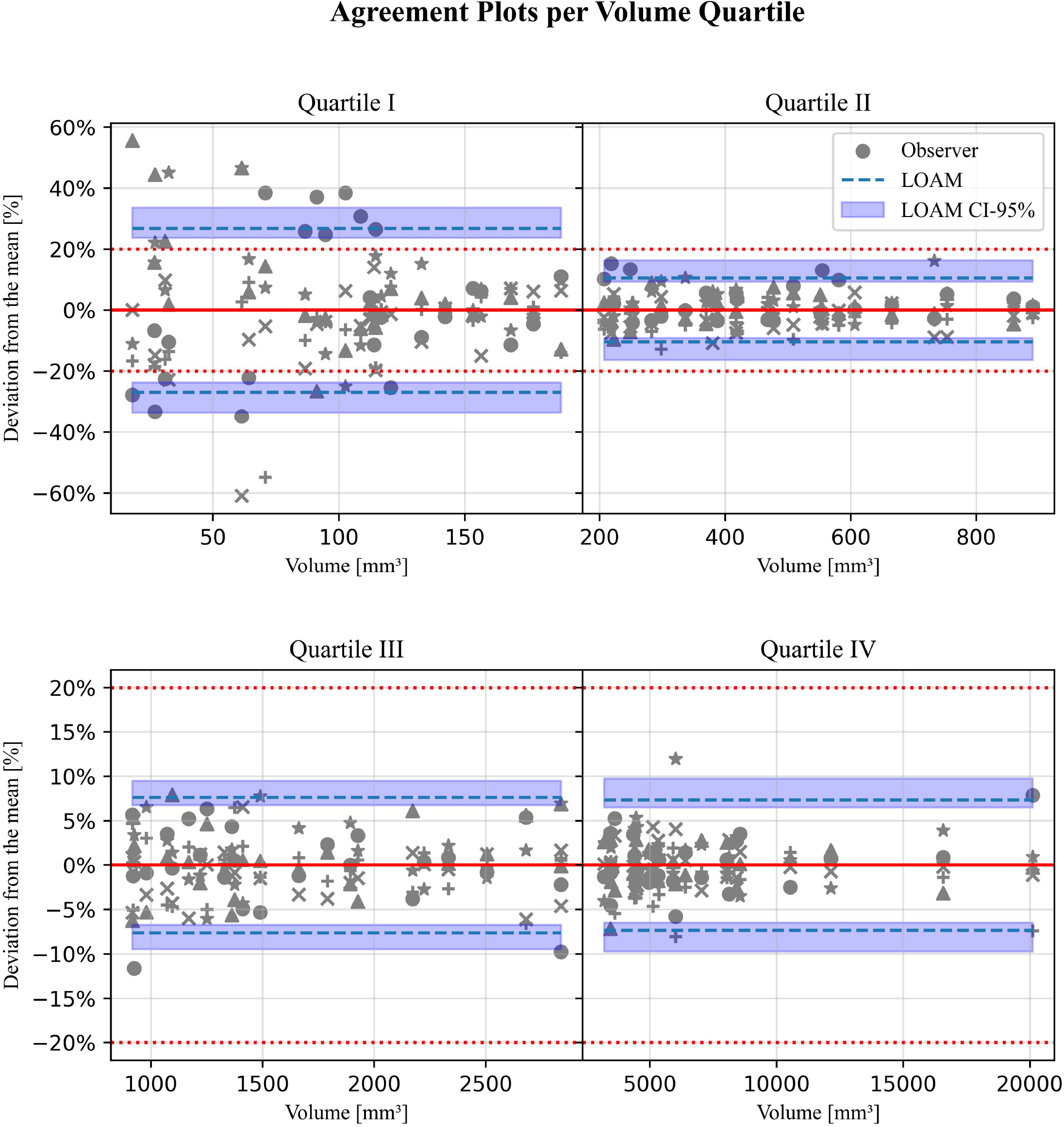
Agreement plots for 100 samples stratified along tumor volume quartile. The different grey colored shapes represent the 5 different observers. The horizontal red dashed line indicates the current used limit of agreement of 20%.

From these tables and figures, it can be discerned that the LOAM relative to the mean (LOAM%) decreases from 26.8% (95% CI: 23.7, 33.6) to 7.3% (95% CI: 6.5, 9.7) for increasing size categories I to IV. This is in line with the calculated ICC values, since the reliability of the annotations increases with the tumor size category (from 0.911 [95% CI: 0.849, 0.954] to 0.995 [95% CI: 0.991, 0.999]). For all four categories, the ICC values show excellent agreement, i.e. higher than 0.9^20^.

When tumors with peripheral cysts are excluded, a similar pattern of volume dependency can be observed. For size category IV, the exclusion of these tumors results in a significantly lower LOAM% (4.8% [95% CI: 4.2, 6.2] cf. 7.3% [95% CI: 6.5, 9.7]), where the CIs do not overlap.

The LOAM method allows for the estimation of different components in the variance between annotations, i.e. an estimation of the source of the variance. The estimated inter-observer variance 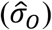 is low compared to the mean value. This is further reflected in the determined high ICC values: there is an excellent (ICC>0.9) agreement with the annotations. The estimated residual variance 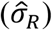 is higher than the inter-observer variance 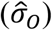 for all size categories. This variance component is related to the extent of the repeatability of the annotations^21^. When tumors with peritumoral cysts are excluded, observers show considerably lower residual variance in size category IV (170 mm^3^ [95% CI: 147, 202] vs. 282 mm^3^ [95% CI: 247, 328]) with no overlap between their CIs.

To illustrate some of the differences in annotations, Figure 3 shows the images of tumors with the highest relative variance between observers for each of the four size categories, along with the two annotations that deviated the most.

**Figure 3:**
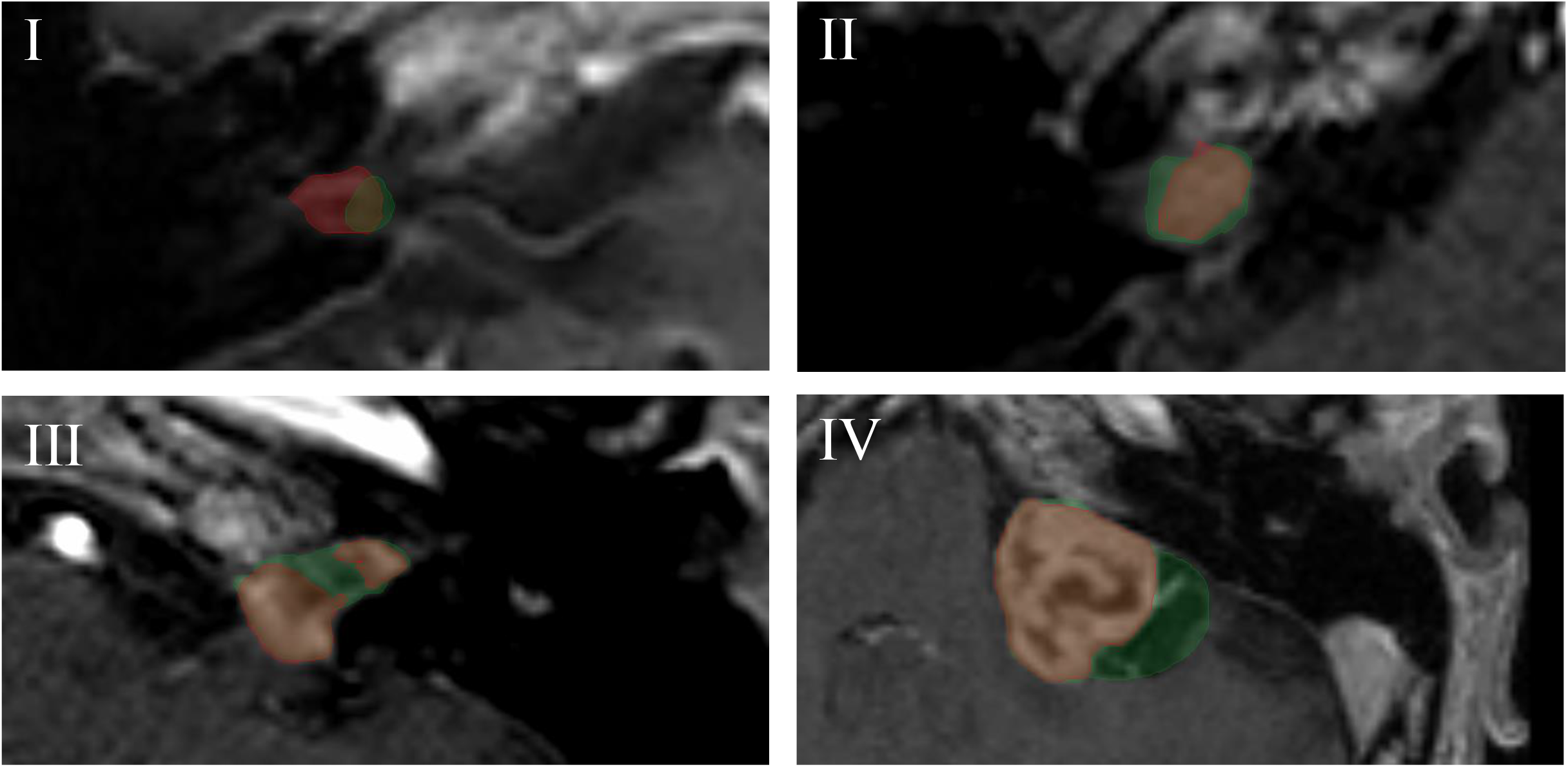
Contrast-enhanced T1-weighted images of the cerebellopontine region on the axial plane. Images correspond to the subjects with the highest relative variance between observers for each of the four size categories. Within the image, the shown annotations (red and green) are the ones that deviated the most from each other. (I and II) Intracanalicular VSs with low signal intensity, partially due to the partial volume effect. (III) VS with a concave surface towards the inferior direction, which causes low signal intensity in the center part of the tumor. (IV) VS showing thin walled peritumoral cysts.

### 3.3 Overall agreement limits

The agreement plot corresponding to the continuous volume-dependent LOAM for all tumors is presented in Figure 4. Herein, the earlier observed negative relation between volume and LOAM% is evident. A steep drop in the agreement limits for lower volumes is followed by a more gradual decline after which the LOAM% appears to stagnate towards a fixed lower limit. The upper bound of the CI intersects the currently widely applied 20%-agreement limit line at a volume of 200 mm^3^.

**Figure 4:**
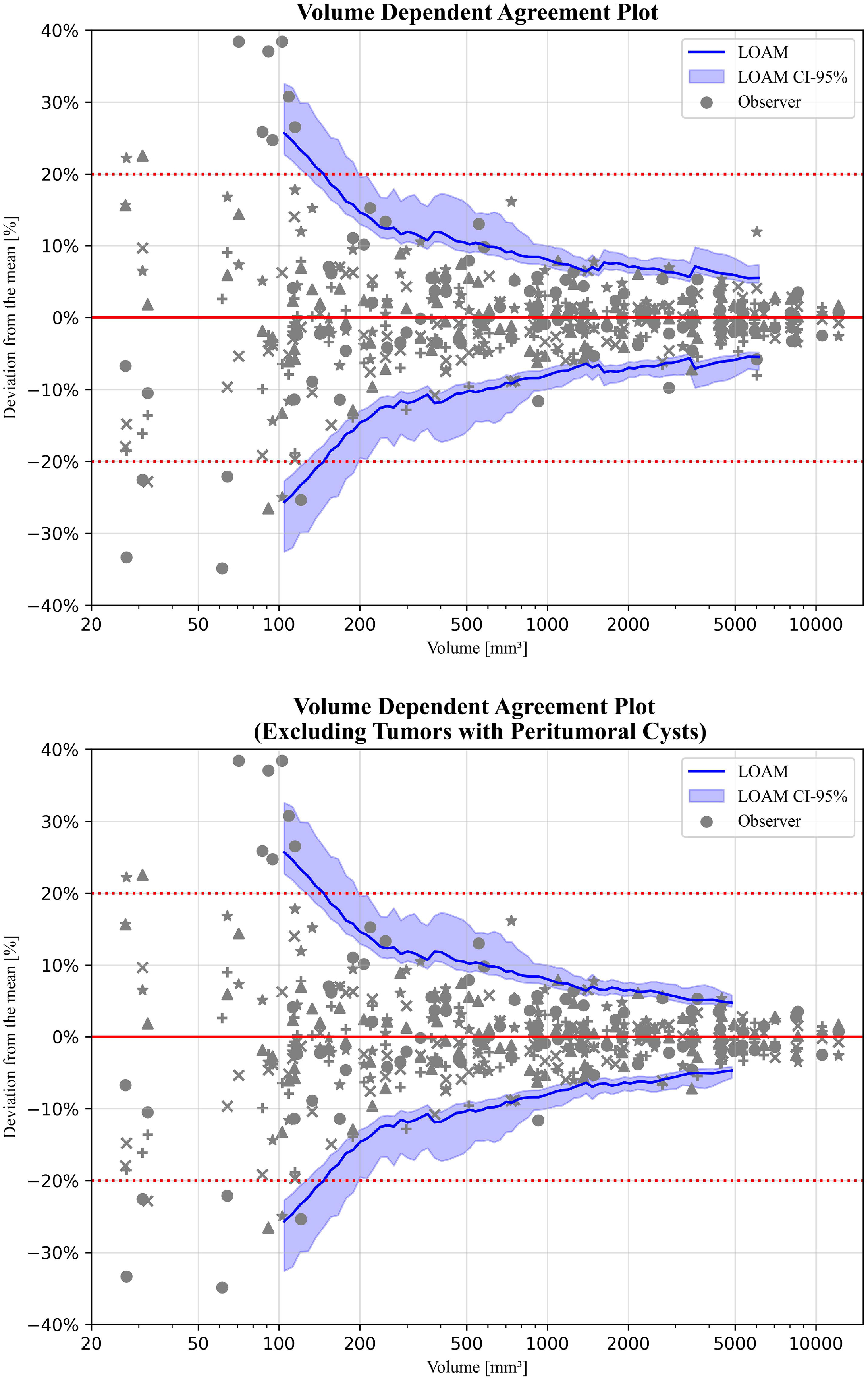
Agreement plots using a sliding window for: (upper) 100 samples stratified along tumor volume quartile, and (lower) 94 samples stratified along tumor volume quartile and excluding cystic tumors. The different grey colored shapes represent the 5 different observers. The horizontal red dashed line indicates the current used limit of agreement of 20%.

When considering individual annotations, not all tumors in the low-volume range exhibit a high degree of variance in their annotations. This is reflected in the wide CI for smaller tumors.

Conversely, for larger tumors, the CI narrows when the number of annotations that highly deviate from the mean decreases. However, a sudden increase in the LOAM% and widening of its CI can be observed at around a volume of 3500 mm^3^. This increment is absent when tumors with peripheral cysts are excluded.

### 3.4 Influence of imaging parameters and tumor characteristics

The results of the univariable and multivariable linear regression analyses are presented in Table 3. Several variables in univariable analysis display a significant correlation with the relative volume standard deviation *SD*_*V*%_. For the imaging parameters, these are: the echo time (B=1.3 [95% CI: 0.0, 2.5; P=0.04]), the magnetic field strength (B=-3.7 [95% CI: -7.4, 0.0; P=0.048]), the slice thickness (B=6.1 [95% CI: 1.4, 10.8; P=0.01]), and the spacing between slices (B=11.9 [95% CI: 1.8, 22.0; P=0.02]). For the tumor characteristics, only the volume is a statistically significant variable (B=-0.001 [95% CI: -0.001, 0.000; P<0.001]). To determine possible confounding effects of the other covariates, a multivariable linear regression was conducted for the statistically significant variables in univariable analysis. Hereafter, only the volume remains associated to the relative volume standard deviation *SD*_*V*%_ (adjusted B=-0.001 [95% CI: -0.001, 0.000; P=0.003]).

**Table 3.**
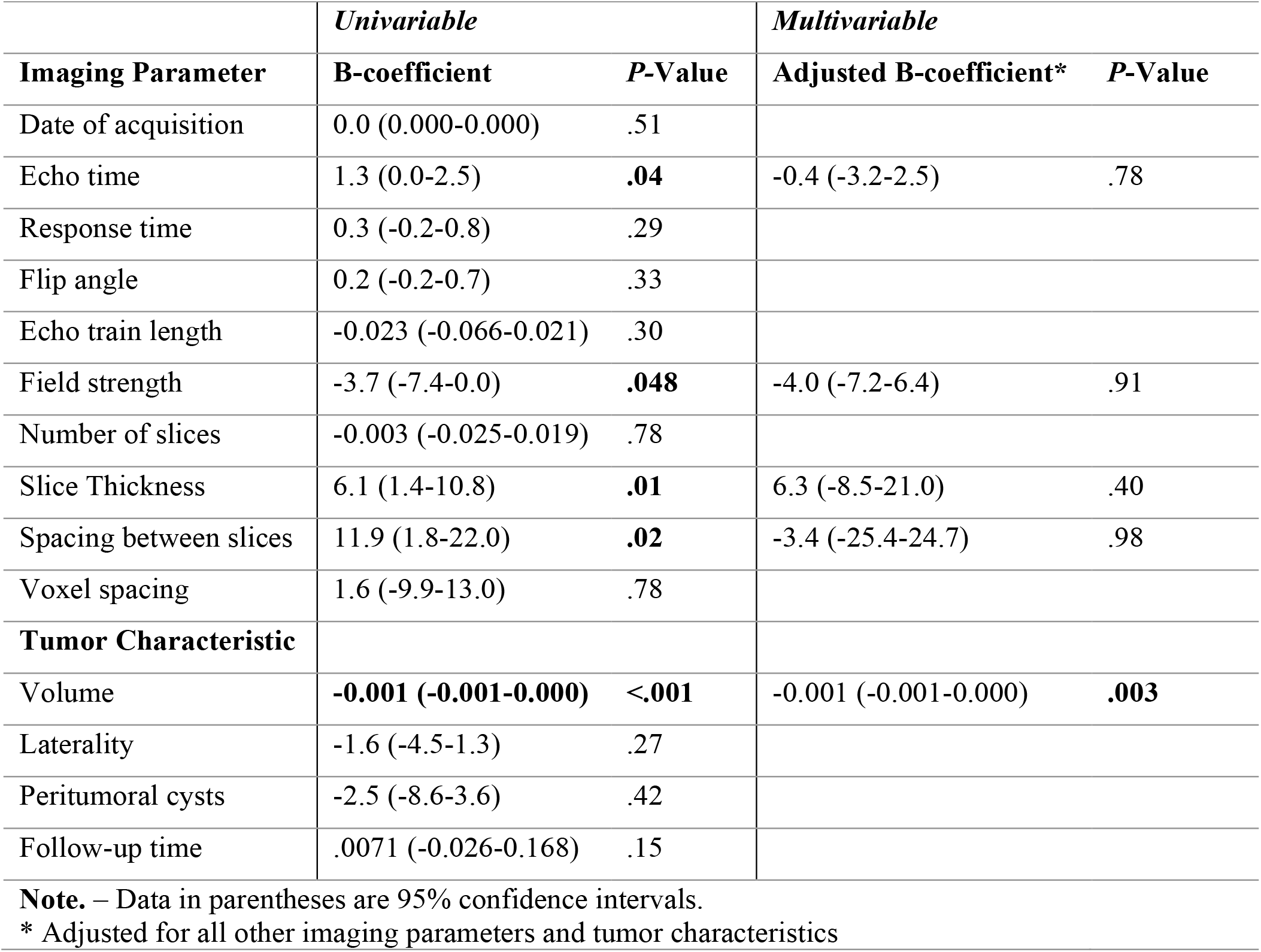
Uni- and multivariable linear regression analysis of imaging parameters and tumor characteristics for the relative volume standard deviation (*SD*_*V*%_)

| Imaging Parameter | *Univariable* |  | *Multivariable* |  |
| --- | --- | --- | --- | --- |
|  | B-coefficient | P-Value | Adjusted B-coefficient* | P-Value |
| Date of acquisition | 0.0 (0.000-0.000) | .51 |  |  |
| Echo time | 1.3 (0.0-2.5) | **.04** | -0.4 (-3.2-2.5) | .78 |
| Response time | 0.3 (-0.2-0.8) | .29 |  |  |
| Flip angle | 0.2 (-0.2-0.7) | .33 |  |  |
| Echo train length | -0.023 (-0.066-0.021) | .30 |  |  |
| Field strength | -3.7 (-7.4-0.0) | **.048** | -4.0 (-7.2-6.4) | .91 |
| Number of slices | -0.003 (-0.025-0.019) | .78 |  |  |
| Slice Thickness | 6.1 (1.4-10.8) | **.01** | 6.3 (-8.5-21.0) | .40 |
| Spacing between slices | 11.9 (1.8-22.0) | **.02** | -3.4 (-25.4-24.7) | .98 |
| Voxel spacing | 1.6 (-9.9-13.0) | .78 |  |  |
| **Tumor Characteristic** |  |  |  |  |
| Volume | **-0.001 (-0.001-0.000)** | **<.001** | -0.001 (-0.001-0.000) | **.003** |
| Laterality | -1.6 (-4.5-1.3) | .27 |  |  |
| Peritumoral cysts | -2.5 (-8.6-3.6) | .42 |  |  |
| Follow-up time | .0071 (-0.026-0.168) | .15 |  |  |

**Note.** – Data in parentheses are 95% confidence intervals.
\* Adjusted for all other imaging parameters and tumor characteristics

## 4 Discussion

In current clinical practice, linear measurements of VS remain commonplace, while multiple studies have shown that volumetric measurements are more sensitive^9–13^. Volumetry will conceivably become more prevalent in the near future due to automatic segmentation algorithms powered by artificial intelligence^22,23^. The accuracy and reliability of tumor volume monitoring is subject to variability between observers. To overcome this issue, the current consensus within VS management professes using a cutoff of 20% change in tumor volume to define tumor growth^2,11,12,15^. However, this threshold may not be suitable for all tumors, because the inter-observer variability is considered to depend on volume^11^. To investigate this dependency, we conducted an inter-observer variability study with five observers and 100 unique patients with unilateral sporadic VS, covering a wide range of tumor volumes.

The LOAM method was used to estimate the LoA^19^. When stratifying the tumors across volume quartiles (I to IV), we found the LOAM to decrease with increasing tumor size category. The LOAM% was found to be as high as 26.8% for the smallest tumor quartile and as low as 4.8% for the largest tumor quartile, when excluding tumors with peritumoral cysts. Within the sliding window LOAM, we further observed that the upper bound of the CI intersects the currently widely used 20% standard at a tumor volume of around 200 mm^3^. This indicates that for tumors smaller than 200 mm^3^ the agreement limits are higher than 20% and growth can therefore only be assessed reliably when a higher cutoff is used. On the contrary, growth in tumors larger than 200 mm^3^ can reliably be assessed with a lower cutoff. Additionally, both univariable and multivariable linear regression showed highly significant negative correlations between volume and the relative volume standard deviation *SD*_*V*%_.

This all provides evidence to our hypothesis that inter-observer variability is dominantly dependent on tumor volume. Therefore, the currently widely used static definition of 20% as a cutoff is not suitable to reliably assess tumor growth, since this will most likely result in either an overestimation or underestimation of the number of growing tumors for tumors smaller or larger than 200 mm^3^, respectively. We created an online calculator (https://vs-study.shinyapps.io/loamcalculation/), where individual tumor volume data can be entered to calculate the specific LOAM% with its corresponding CI.

There is a considerable disparity in the random measurement error (i.e. the estimated residual variance) of all tumors compared to tumors without peripheral cysts for size category IV. The exclusion of these cystic tumors causes a decrease in residual variance, which indicates that the presence of peritumoral cystic components results in a lower degree of annotation repeatability^21^. This is further exemplified by the discrepancy between the two annotations for tumor IV in Figure 4. Herein, one annotator overlooks the thin-walled peritumoral cystic component of the tumor. An explanation for this may be the similar low signal intensity that a peritumoral cyst has, compared to the cerebral spinal fluid on a contrast-enhanced T1-weighted MRI.

We found no significant association between the imaging parameters and the relative volume standard deviation *SD*_*V*%_ in our multivariable analysis. The imaging parameters that showed statistical significance in the univariable analysis can be explained by the uneven distribution of tumor volumes across parameters. For instance, patients who underwent a 3.0 T MRI all had large tumors. However, the results of the multivariable analysis suggest that the quality or type of the image has no relevant effect on the variation of annotations between observers in this study.

The findings in this study provide evidence that volumetric measurements should be interpreted differently than previously thought in order to improve its robustness and reliability. The volume-dependent LoA and therewith definition of growth provides a tailored metric for each individual patient. Tumor response can thereby be monitored more accurately, which is relevant to clinical decision making after radiosurgery or within a wait-and-scan management strategy. However, it should be noted that a difference exists between measurable growth, as presented in this study, and clinical relevant growth. A volumetric increase of 20% for a small tumor is clinically less relevant than the same increase for a large tumor where significantly more adverse effects would arise^12^. Therefore, the ability to reliably detect (measurable) growth in large tumors in an earlier stage will benefit individual clinical decision making. Furthermore, a more reliable and uniform definition of volumetric tumor response will benefit scientific research. Reports of tumor response within patient cohorts can be described with more accuracy, which in turn will improve analyses on treatment efficacies. Finally, it may be possible to extend this concept of volume dependency to other, radiologically similar contrast-enhancing intracranial lesions (e.g., meningiomas, and other intracranial schwannomas).

This study is subject to some limitations. Firstly, even though no relationship was found between imaging parameters and inter-observer variability, the single-center nature of this study exposed the observers to a more homogeneous image dataset than in a multi-center setting. An earlier study on potential error sources for tumor volumetry, for instance, found a negative effect of a large slice thickness^24^. This effect was not present in our study. As such, the LoA and thereby the growth definition cutoff is plausibly higher in data that is more heterogenous. Nevertheless, we conjecture that our findings on the volume-dependency of the agreement limits will still hold. Secondly, our study focuses exclusively on contrast-enhanced T1-weighted MRI. Other image sequences may result in different agreement limits, which is demonstrated in an earlier study on T2-weighted images^11^. Our results are therefore only applicable to volumetry on contrast-enhanced T1-weighted images.

## 5 Conclusions

We have shown that inter-observer variability in volumetric VS annotation is mainly affected by tumor volume. The currently widely used approach to define tumor growth in VS, i.e. 20% change in volume, should therefore be reconsidered. In most cases, tumor growth can reliably be detected at an earlier stage due to the lower found limits of agreement. However, caution should be exercised for small tumors, i.e. smaller than 200 mm^3^, as agreement limits are higher than previously thought.

## Data Availability

All data produced in the present study will not be publicly available

